# Evaluation on the Effectiveness on the Implementation of WHO-CST Programme in Hong Kong: A RCT Protocol

**DOI:** 10.1101/2021.05.17.21257327

**Authors:** Paul Wai-Ching Wong, Siu-Lun Chow

## Abstract

Developmental delays/disorders in young children are identified as a public health priority. WHO and Autism Speaks co-designed a training programme titled “WHO-Caregiver Skills Training” (WHO-CST) which aims at providing training for caregivers whose children are with possible symptoms of Autism spectrum disorder (ASD) symptoms and pervasive developmental disorders (PDD), so that they can learn better strategies on managing the daily lives of their children and foster better communication between the parent-children dyad. The WHO-CST programme is currently implemented in more than 30 regions worldwide and it started in Hong Kong from 2018. As the programme is newly developed, only a few studies tried to evaluate the effectiveness of the programme in qualitative approaches, or in quantitative approach with relatively small sample (e.g. n < 10). In the present study, our team, who is responsible for the implementation and evaluation of the WHO-CST programme in Hong Kong, attempt to assess the effectiveness of the training in WHO-CST under a randomized controlled trial (RCT) design with about 120 eligible caregivers who will be randomly assigned in experimental and control groups. Our measurement will include the Joint Engagement Rating Inventory (JERI) developed by experts in WHO to gauge how parents engage their children in a 10-minute video recording their dyadic interaction and behaviors in a defined play setting. A set of other measurements on the caregiver’s experience of using intervention skills and their enhancement of knowledges will also be measured. We hypothesize that caregivers in treatment group will have better growth in scores of both JERI and measurements of other outcome than the wait-list control group’s after the 12-week WHO-CST training, and also persistence of skill and knowledge level should also be found after a 30-day follow-up test.

## Introduction

The DSM-5 defined Autism spectrum disorder (ASD) as persistent deficits in social communication and social interaction across multiple contexts, but in clinical context, it usually refers to a set of various neurodevelopment disorders, which can possibly include autism, Asperger syndrome, pervasive developmental disorder not otherwise specified (PDD-NOS), and other similar conditions (Qiu et al., 2020). Survey figures of diagnosed ASD cases have been raised worldwide in the last two decades. The Autism and Developmental Disabilities Monitoring (ADDM) Network estimated the prevalence of ASD among children aged 8 years was 18.5 per 1,000 children in eleven selected states in America in 2016, while other recent reports from the ADDM network stated that in United States, the 4-year-old ASD prevalence and the 8-year-old ASD prevalence were 13.4 per 1,000 (Christensen et al., 2016), and 16.8 per 1,000, respectively (Baio et al., 2018). These figures have been doubled when compared to those reported in the past 10 years (i.e., 6.7 per 1000) (Baio et al., 2018). A recent meta-analysis in 2020 found that the pooled estimate of ASD prevalence was 0.36% in nine selected Asian countries3 which was much higher than that in similar analysis in 2010 (0.15%) (Qiu et al., 2020), and the pooled estimate of ASD prevalence in mainland China was about 0.39% in a meta-analysis at 2018 (Wang et al., 2018).

Taking care of children who have ASD symptoms can be difficult, especially when they may exhibit one or more of the core symptoms, such as impairments in social interactions, communication and behaviour malfunctioning (Johnson & Myers, 2007). Many children having ASD children were found co-occurring intellectual disability (intelligence quotient [IQ] ≤70) (Maenner et al., 2020), or behavioural problems, such as noncompliance, aggression, and self-injury (Soke, Maenner, Christensen, Kurzius-Spencer, & Schieve, 2018). As children with ASD symptoms usually have limited capacities of acquiring daily living skills, many parents face challenges in parenting, which can severely impact family functioning and their psychological well-being (Montes & Halterman, 2007). This can create stress to caregivers since taking care their children who have ASD may reduce their own ability to socialize with other and further bring negative impact on the relationships with their spouses or partners (Sim, 2018). In the context of atypical neurodevelopment, ASD and intellectual disabilities can be associated with altered parental responding and directiveness towards the child, possibly due to parents’ difficulty in interpreting child’s behaviour accurately or because the child has poorly regulated interaction and attention (Green & Garg, 2018).

### Children with ASD in Hong Kong

In the case of Hong Kong, there was no official estimation on the number of ASD prevalence in the whole population, but several related statistics could be retrieved from reports of different organizations and government departments. For instance, number of students with autism spectrum disorders in mainstream public schools rose from 2,050 in 2010, to 8,710 in 2018 (Sun, 2020). In primary schools, 24% of 22,980 students who have special needs were diagnosed as having ASD in 2018 (Legislative Council Secretariat, HKSAR, 2019). With reference to the latest Mental Health Review Report (Food and Health Bureau, HKSAR, 2017), the caseload of Child and Adolescent Psychiatric Services under the Hospital Authority for ASD alone increased from about 5,000 in 2011/12 to about 9,000 in 2015/16, comprised over 60% of the caseload of the Services. The number of children newly diagnosed with ASD in the Child Assessment Service of the Department of Health has increased close to three times from 755 in 2006 to 2021 in 2015, the number of preschoolers with significant developmental disorders has also been doubled (Lee, 2016). All abovementioned evidence showed that the figures of children having been diagnosed as having ASD in Hong Kong have been soared rapidly in the last two decades.

Living in a Chinese society like Hong Kong, parents of children with ASD diagnosed have to face strong pressure from the society since the traditional Chinese culture, which always emphasizes the association of parent’s competence with children’s academic achievement, reinforces discrimination on having a child with disability (Mak & Kowk, 2010). Parents tended to internalize such external criticism (as affiliate stigma) on their children, and assume their own responsibility on the stigmatic condition, and believe that they may be utterly uncontrollable to their children’s condition and corresponding stigmatization. Such affiliate stigma brings psychological and parenting stress to the parents, and consequentially damages their mental health and psychological well-being (Wong, Mak, Liao, 2016; Chan & Lam, 2018), delays formal diagnosis as a denial to children’s ASD symptoms, and sometimes hinders their children’s involvement in community participation (Ng et al, 2020).

Cost of available private services (including diagnosis) for children with ASD symptoms was high and less affordable by lower class in Hong Kong (Mak & Kowk, 2010; Tait et al., 2016). Meanwhile, although the government provides lower-cost public services such as early diagnosis and medical treatment through Child Assessment Centres and NGO services at district level for children under age 12, they were not sufficient to fulfill the overwhelming demand, resulted in a rather long waiting period (about 12 to 24 months) for initial developmental assessment (Wong et al., 2015). The delay of diagnosis can bring ramification on children’s school enrollment and procrastinate necessary treatment and services (Yi, Siu & Chan, 2020). Self-stigma, anxiety and hesitation on seeking formal diagnosis and services may also attribute to parent’s lack of awareness and knowledge on ASD and developmental delay. Although Hong Kong has a relatively advanced and well-organized public health system compared with other less developed cities, parents may still not have sufficient knowledge and information on accessing clinical professionals and services at their living districts (Ho, Yi, Griffiths, & Murray, 2014; Yi, Siu & Chan, 2020).

Therefore, at the time being for parents who suspects that their children may have ASD are waiting for receiving formal diagnosis, pre-assessment intervention can be offered to enhance their caregiving skill and knowledge on handling situation of ASD children. Services or interventions which allow parents to communicate with professionals and other parents having similar caregiving experience can also moderate their anxiety, since social support and teaching parents to have positive perception can ameliorate mental and relational well-being among parents of children having ASD (Koegel, Koegel, Ashbaugh & Bradshaw, 2014; Wong, Mak, Liao, 2016). Well organized pre-assessment service can also be associated with a reduced duration of diagnostic process for children. Offering adequate and relevant information in pre-assessment period can be likely to reduce total duration of the assessment process (McKenzie, 2015). Recent review on services prepared for ASD children also suggested that provision of pre-assessment information workshops and earlier intervention to parents can reduce their anxiety and let them become more well-prepared for formal diagnosis and services (Ho, Yi,Griffiths, & Murray,2020).

Furthermore, there is growing evidence supporting that enhancement in parental skills in communication, engagement and mitigating autistic mannerisms can be achieved through appropriate intervention, leading to better developmental and behavioural outcomes of ASD children, and also better family functioning (Oono et al., 2013; Reichow et al., 2013). Involving parents in implementing interventions to their children allows consistent handling, and ensures that the intervention is appropriate in enhancing child’s earliest social and communicative development (Green & Garg, 2018).

Since August 2015, the JC A-Connect Family Support Team (hereinafter, “the Team”) was set up to provide early pre-assessment intervention and various training programmes for parents who suspected that their children may have ASD. The team was funded by the Hong Kong Jockey Club Charities Trust and wholly managed under the Faculty of Social Sciences, the University of Hong Kong, and the programmes launched were co-operated with several NGOs in Hong Kong (i.e., Caritas-HK, Heep Hong Society, and SAHK). A few small-scale evaluative studies have been conducted to examine the efficacy of different programmes, and these studies showed small within-group effect sizes on the reductions in child’s problematic behaviours, parenting stress, as well as on the improvements of perceived social support, parenting competence and mental health (Cohen’s *d* ranged 0.11-0.22; 226 families). While the Team appreciated the creativity and commitment of the NGOs in designing innovative strategies to address the needs of families of children with ASD, the non-standardized programme implementation and non-comparable evaluation design resulted in difficulties on assessing the successfulness of the programmes and failed to inform any measures on improving these set of programmes. Under consideration of these defaults, at the beginning of 2019, the Team began to adapt a standardized training programme titled “World Health Organization - Caregiver Skills Training” (WHO-CST) in Hong Kong, which was co-created and recommended by WHO and Autism Speaks. Eight major NGOs (Caritas Hong Kong, Heep Hong Society, SAHK, the Salvatory Army, the Tung Wah Group of Hospitals, the Hong Kong Sheng Kung Hui Welfare Council, YWCA, and the New Life) which have provided services for children with ASD also joined the project and implemented the WHO-CST programme in various districts of Hong Kong under the coordination of the Team.

### The WHO-CST Programme

Developmental delays/disorders in young children are identified as a public health priority. In 2009, the World Health Organization (WHO) launched the mental health Gap Action Programme (mhGAP) aiming at bridging up the treatment gaps for mental, neurological and substance use disorders worldwide (World Health Organization, 2009). In the latest mhGAP intervention guide, WHO and Autism Speaks co-created and recommended that the Caregiver Skills Training (CST) programme should be widely implemented in the management of children with possible ASD symptoms, intellectual disabilities and pervasive developmental disorders (World Health Organization, 2016). The WHO-CST materials were developed based on findings of several meta-analyses (Oono et al., 2013; Reichow et al., 2014; Reichow et al., 2013) and in consultation with experts and parents’ association from all WHO regions and support from Autism Speaks (World Health Organization, 2017). The aim of the WHO-CST programme is, on one hand, to train non-specialist social service providers, such as social workers

The WHO-CST programme package, hence, includes the following characteristics:

- a multi-component intervention in which its content is theoretically based on behavioural approaches for promoting shared engagement and communication, such as Joint Attention Symbolic Play Engagement Regulation (JASPER), Pivotal Response Treatment (PRT) and Discrete Trial Training (DTT), as well as positive parenting approaches for promoting positive child behaviour, and/ or management of challenging behaviour;
- a programme consisted of nine group sessions and three individual home visits, focusing on training parents on how to use every day play and home activities and routines as opportunities for learning and development;
- a programme required vigorous fidelity monitoring (e.g., video-recording facilitators’ performance) and comprehensive evaluation on both parental and child outcomes by self-reports and through blinded, trained assessors;
- a task-shifting approach in which non-specialists, such as social workers, teachers and community leaders, can train parents to deliver interventions for promoting their child’s development effectively; and
- a trans-diagnostic approach, so that children who have not met the diagnostic criteria for ASD or other pervasive developmental disorders (such as developmental delays) could be benefited from the programme.

Under the framework of implementation science, the WHO-CST programme was in the stage of pilot-testing in more than 30 countries at 2018 (World Health Organization, 2018) and several randomized controlled trial was underway in Pakistan (Hamdani et al., 2017) and other countries.

In the actual implementation, the WHO-CST programme consisted of 12 sessions, which was comprised of:

1. 9 weekly group-based sessions (each session will last approximately for 2.5 to 3 hours), 6 to 8 caregivers in a group, led by 2 facilitators who have received a 5-day training by WHO staffs and Autism Speaks, and,
2. 3 home visits (each visit will last approximately for 1 to 1.5 hours) occurred before the 1st session, between session 4 and 5, and after the last session, conducted by 2 facilitators to each caregiver’s family.

The themes and detailed objectives of each session were listed in Table 1.

**Table 1.**
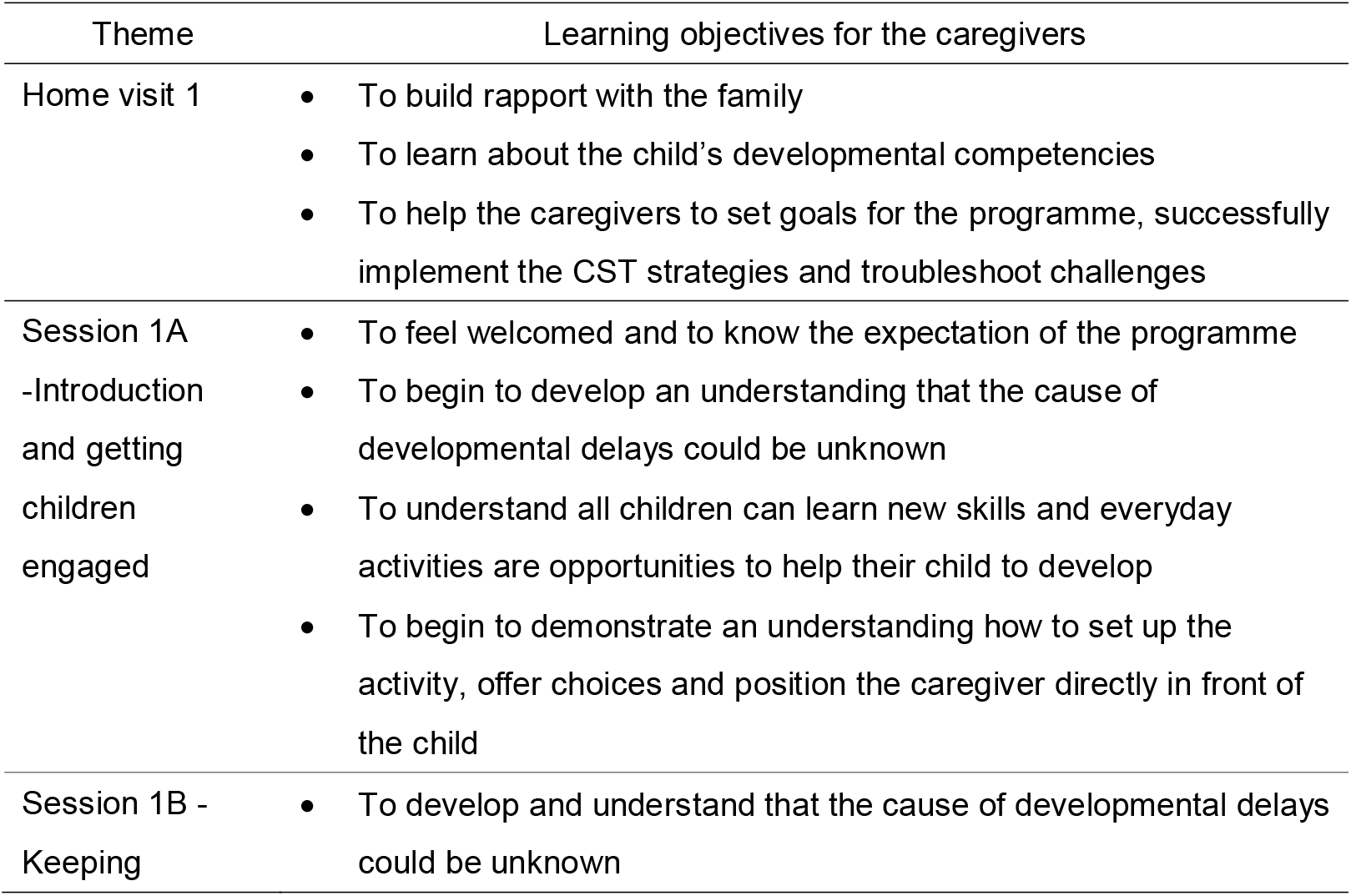
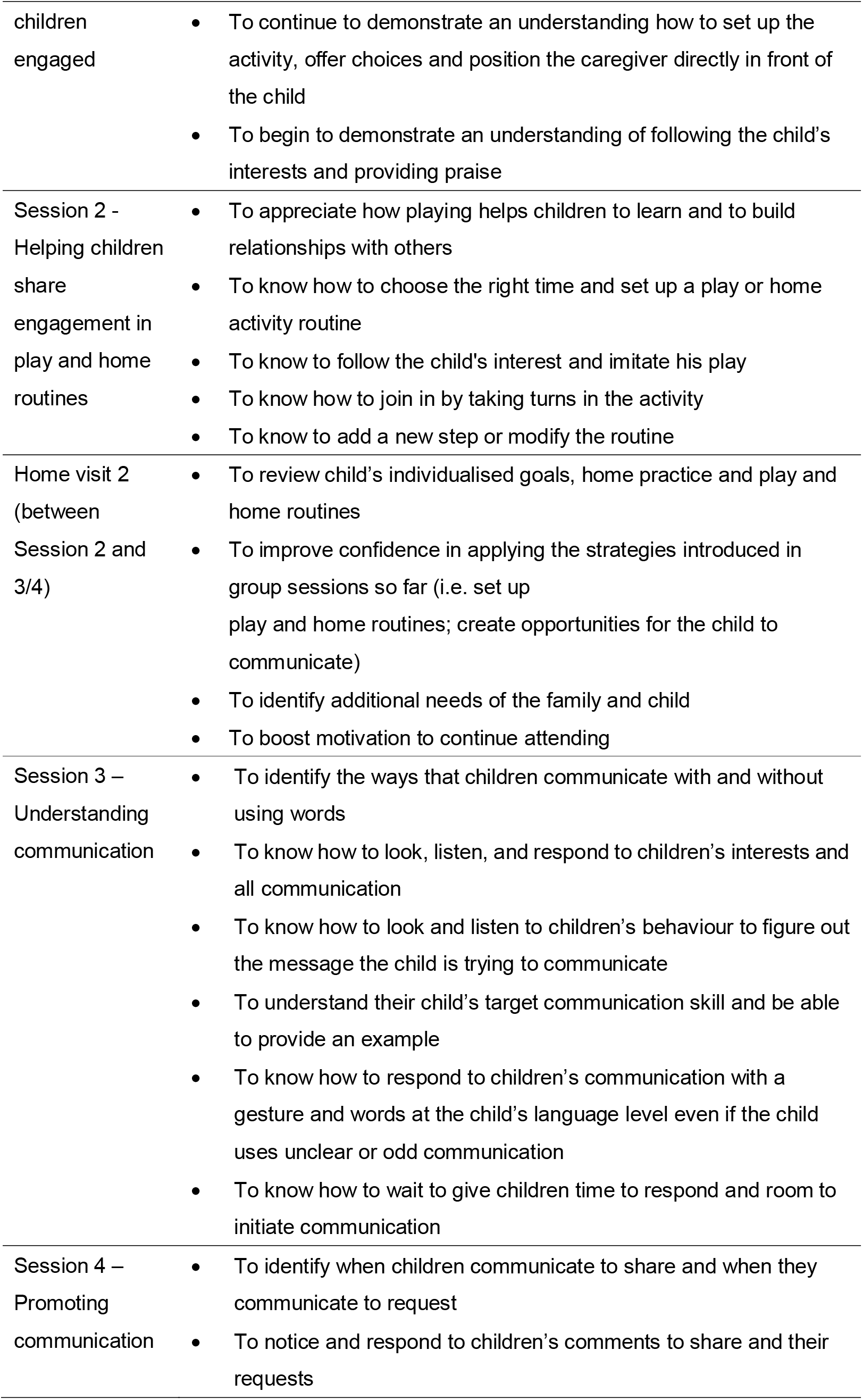
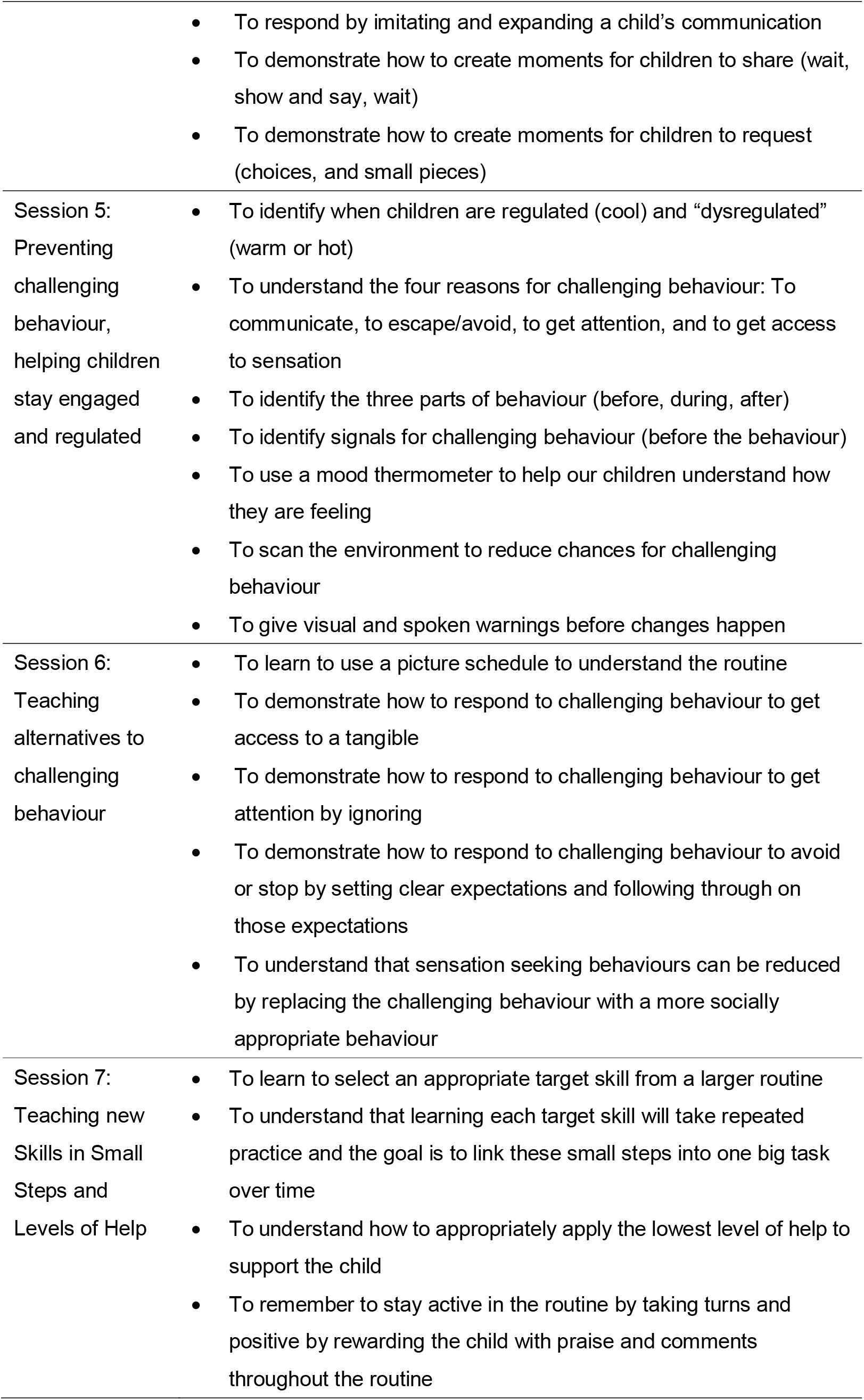
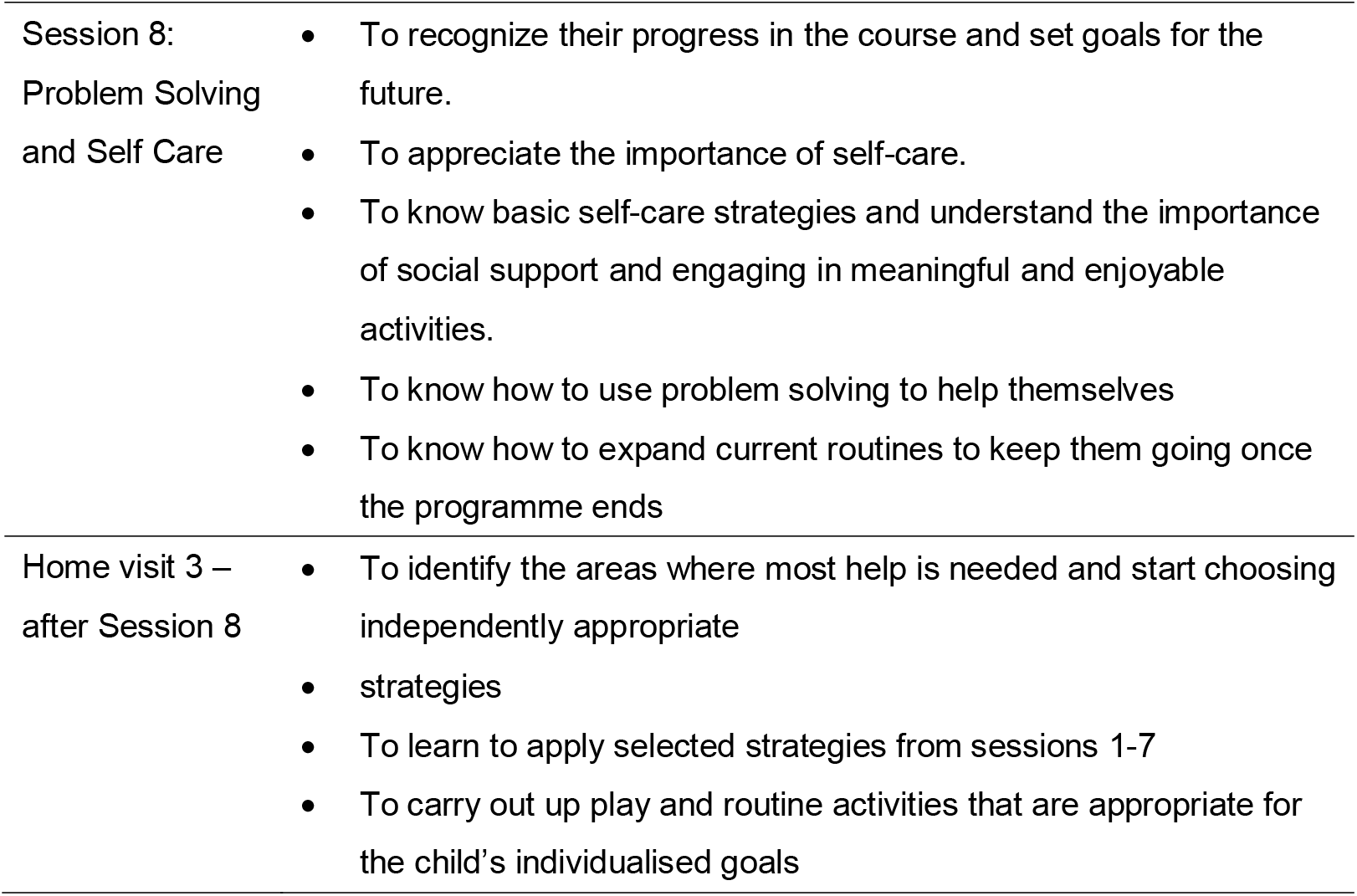
Session-by-session outline of the WHO-CST programme

The group-based sessions in delivering CST materials focus on training caregivers to improve communication skills, shared engagement in activities and routines with caregiver-child dyads, behavioural skills and problem-solving ability in caring for a child with developmental delays and/or disorders. Each group session consists of the following key activities:

- Brief wellness activity, e.g., abdominal breathing exercise
- Review of key CST messages and home practice
- Sharing of a case scenario related to the child’s developmental difficulties
- Group discussion and experience sharing
- Skills demonstration by facilitators
- Live practice among caregivers in pairs
- Establish plan for home practice and session review

Home visits provide an opportunity to build rapport with the family, learn about the child’s developmental competencies and behaviour and the home family environment, help the caregivers to set goals for the programme, successfully implement the strategies that have been introduced to during the group sessions, troubleshoot challenges that have been encountered by caregivers and identify any additional needs to the family.

Social workers or clinical psychologists had to receive training from WHO experts and passed tests assessing their skills on interacting and communicating with children with ASD symptoms in other to be qualified in teaching the WHO-CST programme. These accredited master trainers were allowed to launch the WHO-CST programme themselves, and train other trainers to implement the programme. On the contrary, the trainers instructed under these accredited master trainers were allowed to deliver the programme to parents, but they were not allowed to train other trainees.

In the case of Hong Kong, all teaching materials in the WHO-CST programme were translated into Traditional Chinese. Lessons and guidance in the home visit were all conveyed in Cantonese, which is the colloquial language used by the major population in Hong Kong. During the COVID-19 pandemic from the beginning of 2020, all the group-based sessions were delivered in the way of video lectures and tutorials (e.g. small-group discussions) through ZOOM, and the home visit session, which investigated how parents interacted with their children, were taken place in training room situated in the University of Hong Kong.

## The Present Study

This study aims to, first, evaluate the usefulness of the WHO-CST intervention in the Chinese-based community of Hong Kong through a randomized controlled trial (RCT) design, and second, examine potential factors hindered or promoted the usefulness of the WHO-CST programme.

### Participants

Caregiver-child dyads as potential participants will be recruited through advertisement on social media and promotion at district level through cooperating NGOs. The recruited caregiver-child dyads have to fulfill a set of inclusion criteria in order to be admitted into the WHO-CST programme.

For caregivers, they should be:

- Hong Kong residents
- at least 18 years or above,
- the primary caregiver who is responsible for the role of parenting the child, this could be biological parent (father or mother), guardian or other adult family member (i.e., the same caregiver who will attend the WHO-CST programme if she/he agrees),
- living together with the target child,
- able to communicate in Cantonese,
- able to read and write basic Traditional Chinese, and
- able to stay in Hong Kong for at least six months for most of the time for home visits and 9 sections of face-to-face group intervention and accessible by phone.

In addition, to ensure the adherence of WHO-CST programme, these caregivers shall allow the following procedures to be carried out during the programme implementation:

- agree to take their children to the University of Hong Kong to attend the three home visit sessions and record videos for assessing their interaction and communication with their children
- agree to be recorded by ZOOM in the 9 group-based sections of intervention.

For children who are eligible to participate the WHO-CST programme in Hong Kong, they should be:

- between 2 and 6 years old,
- screened positive on Modified Chinese version of the Checklist for Autism in Toddlers (C-CHAT-23) Wong et al. (2004) or,
- suspected to display symptoms or behaviours of autism spectrum disorder, such as persistent deficits in social communication and social interaction across multiple contexts and restricted, repetitive patterns of behaviours, interest, or activities, or,
- suspected to display symptoms or behaviours of communication disorder, such as persistent difficulties in the acquisition and use of language across modalities and language abilities are substantially and quantifiably below those expected age, or,
- the above symptoms or behaviours are the primary concerns of developmental issues the child face as reported by caregivers or clinicians (children who are suspected to co-morbid other conditions, such as intellectual disability, attention deficit hyperactivity disorder, specific learning disorder and motor disorder are also eligible if these other conditions are not primary concerns of developmental issues)

### Sample Size Estimation

This study targets to test the clinical effect of the treatment on both self-reported outcomes by caregivers and rater-assessed outcomes of caregiver-children interaction and compare these outcomes between the treatment group and wait-list-control group. The Team assumes a conservative effect size estimate of 0.30 (i.e., moderate effect size) for the outcome measures, 80% power at 5% two-tailed significance level; and 10% attrition rate. Based on Karlsson, Engebretsen and Dainty’s (2003) formula and suggestions, 120 caregiver-child dyads are proposed to be recruited and should be representative to draw significant results in the final analysis. Participated caregiver-child dyads will be randomized to the intervention or to a wait list at 1:1 ratio: 60 caregiver-child dyads will be randomly assigned to a treatment group to receive WHO-CST training whereas another 60 caregiver-child dyads will be randomly assigned to a wait-list-control condition. As the final analysis will be done under the intent-to-treat principle, the dropout participants will also be included in the analysis.

### Randomized Control Trial Design

Participated caregiver-child dyad will be enrolled by accredited trainers of the WHO-CST programme based on the selection criteria listed before. Researcher who generates randomized sequence for RCT is independent from the research team who recruits participant. The researcher will put caregivers name into a Python list (in random order, expected 120 caregivers) and use the Python function “sample()” to randomly extract 60 caregivers without replacement. The flow of the RCT design is shown in Figure 1. Before formally starting the WHO-CST intervention, both groups will complete the 1^st^ assessment as the baseline for group comparison. Then, the treatment group will receive 12 weeks of WHO-CST intervention after the screening tasks, while the wait-list-control group will wait for intervention at this 12-week waiting period. The 2^nd^ assessment of both groups come immediately after the completion of treatment group’s intervention. To investigate the persistent effect of the intervention, a follow-up assessment will be implemented 30 days after the completion of the treatment group’s intervention. In other words, the wait-list-control group will take all the three assessments parallelly with the treatment group and will receive the intervention afterwards. Both participants in the treatment group and wait-list-control group will be blind for their assignment to the RCT experiment. The whole RCT is supposed to be completed within seven months, including the intervention to wait-list-control group.

**Figure 1.**
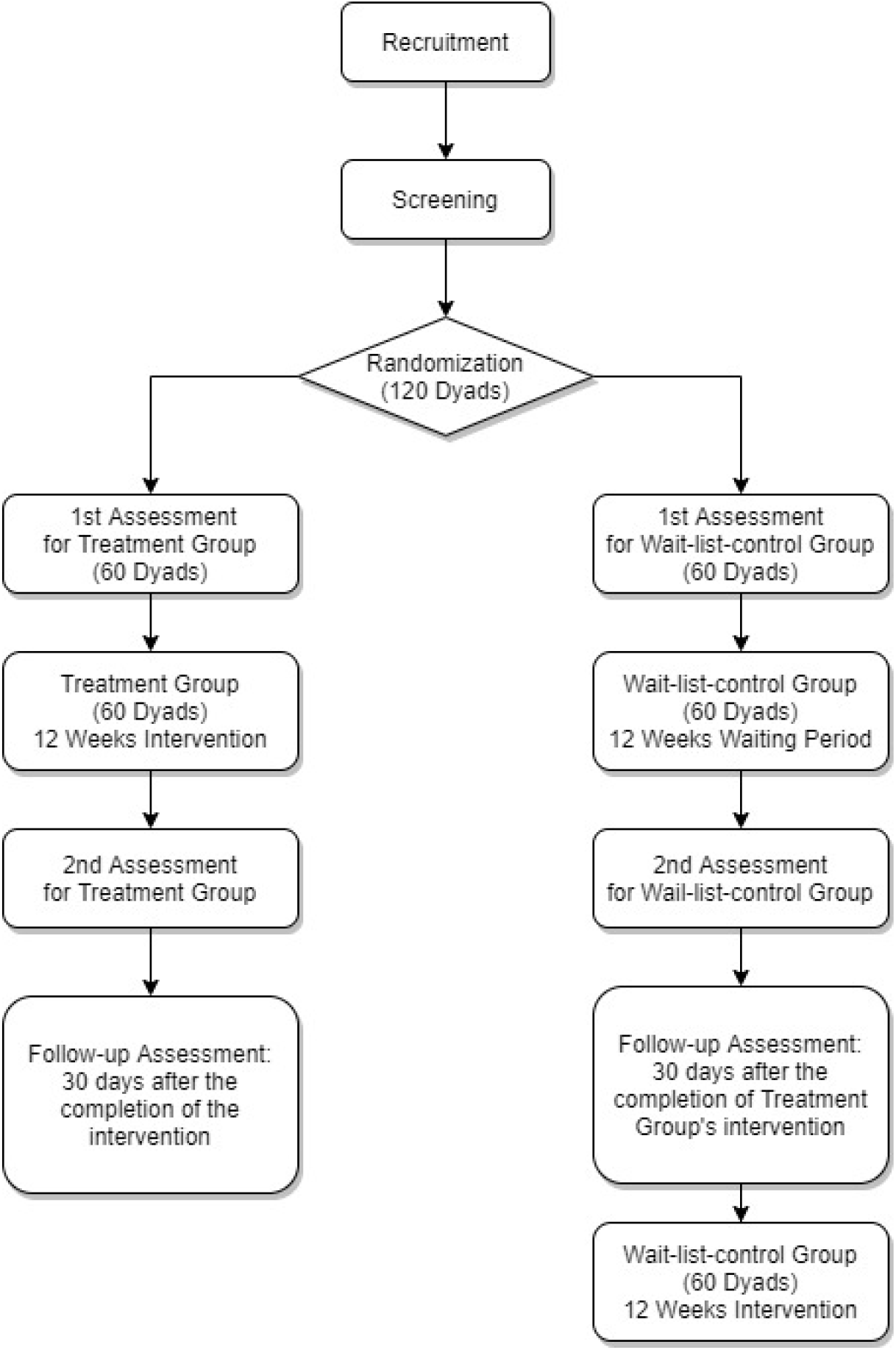
Flow diagram of the RCT design in this study

Since the WHO-CST in Hong Kong is a sustained programme and multiple-parallel training sessions are opened continuously throughout the two years of operation, it is normal that some caregivers have to wait for 3 to 6 months to receive the training. Also, it is a normal practice to have multiple assessments before and during the WHO-CST sessions. The assessment, training content and practices are all identical between the treatment group and the wait-list group. Therefore, the caregivers will only know that they are assigned into different time slots of training, but will not know whether they are in the treatment group or in the wait-list-group.

### Hypothesis

This study hypothesizes that, (1.) after the intervention through the WHO-CST training, treatment group’s caregiving knowledge and skills will be enhanced more than the wait-list-control group and, (2.) after the intervention, the children of treatment group parents will have less misbehavior than those of the wait-list-control group parents.

## Assessment Tools

The assessment tools in this study includes parents’ self-reported on-line survey and video-taking of parent-children playtime in pre-set scenarios in training room at the University of Hong Kong. During the playtime, toys, duration of the playtime and instructions to parents are all standardized to minimize factors of circumstance.

### Measurements in Parents’ Self-Reported Survey

Participants will be invited to complete a questionnaire in soliciting their sociodemographic information and their child’s characteristics at the 1^st^ Assessment. Then, in each round of assessment, both the treatment groups and the wait-list-control groups will be requested to complete the following outcome measures in the self-reported survey. All the outcome measures have been translated into Traditional Chinese.

#### Caregivers’ Knowledge and Skill

The Caregiver Knowledge and Skills Test is designed by WHO expects who led the worldwide WHO-CST programme and will be used to assess the knowledge and skills of the caregivers related to WHO-CST programme materials. Caregivers will be invited to rate 24 statements on a 5-point Likert scale ranging from 1 (I strongly disagree) to 5 (I strongly agree), for example, “My child has more opportunities to learn when we are focusing attention on same toy or activity”. Further, caregivers will be asked to indicate their confidence level (13 statements) on a 5-point Likert scale ranging from 1 (not confident) to 5 (very confident), such as “I feel confident in using pictures to help my child follow a routine”. Caregivers will also be asked to complete 3 scenario-based short answer questions.

#### Caregivers’ Experience on Using Strategies

A set of questions will be used to evaluate the caregivers’ experiences with the intervention strategies that they learnt during the WHO-CST programme, in terms of their level of confidence, comfort in using the strategies, as well as how difficult, effortful and natural the caregiver perceives the strategies (Kasari et al., 2014).

#### Caregivers’ Quality of Life

Parents have to report their quality of life in each assessment through filling in the General Health Questionnaire-12 (GHQ-12). The GHQ-12 is a widely used and easy-to-understand instrument for measuring parents’ psychological strain, particularly in aspects of social dysfunction and anxiety / depression. The Chinese version of GHQ-12 have been well-validated in several studies (Li, Chung, Chui, & Chan, 2009; Wong & O’Driscoll, 2016).

#### Children’s Misbehavior

The suspected ASD children’s misbehaviors will be gauged mainly by two scales. The first one is the Eyberg Child Behavior Inventory (ECBI) which is a 36-item multi-dimensional, parent-rating scale on the perception of their children’s conduct problems, especially for children 2 to 17. Parents are asked to rate how often each stated behavior occurs on a 7-point frequency-of-occurrence scale, which generate the “intensity score”. They are also required to indicate whether the stated behavior is still currently a problem to them, which generate the “problem score”. High scores represent high frequency and high number of children’s disruptive behavior in their daily life. The Chinese version of ECBI have been validated by the Education Bureau (formerly known as the Education and Manpower Bureau), HKSAR (Leung et al., 2005).

The second scale is the Chinese version of the Modified Checklist for Autism in Toddlers (Wong et al., 2004) (Chinese M-CHAT). This 23-item scale is a common and easy-to-administered screening tool for identification of autistic children, which addressed on aspects like children’s social relatedness, joint attention, bringing objects to show parents and their responses to other’s calling. Wong et al. (2004)’s study confirmed the validity of this scale on young children and its reliability for Hong Kong caregivers.

### Measurements on Video-recoded Playtime

For each of the participating parent-dyad, they have to take videos on a standardized 10-minute play scenario at each assessment time (the 1^st^ assessment, the 2^nd^ assessment and the follow-up assessment). All the videos will be rated by the Joint Engagement Rating Inventory (JERI) (Adamson, Bakeman & Sumam, 2020). JERI was designed by experts cooperating the WHO-CST, and was targeted to characterize both child and caregiver’s activities during the Communication Play Protocol (Adamson, 2016). The scale is adapted to rate child’s engagement states as well as various aspects of the children’s and caregiver’s behavior and of their shared activities. Raters of our studies have to view the video records of the 10-minute play scene in the caregiver-child interaction using skills acquired in the WHO-CST training, and they have to make judgment about the interaction using seven-point rating scales on ten items:

1. Unengaged
2. Joint Engagement
3. Stereotyped, Restricted, and Repetitive Behaviours
4. Attention to Caregiver
5. Initiation of Communication
6. Expressive Language Level and Use
7. Scaffolding
8. Following in
9. Caregiver’s Affect
10. Fluency and Connectedness

These ten items address on various aspects of caregiver-child interaction including child’s engagement state (e.g. joint engagement), child’s activities (e.g. initiation of communication), caregiver’s supports on child’s activities (e.g. scaffolding), caregiver’s attention on child’s focus (e.g. following in) and dyadic interaction (e.g. fluency and connectedness).

To maintain fairness and reliability of the rating practices, the rating team will not be members of the research team of this study. The rating team are particularly recruited for rating the videos and have no direct contact nor interaction with any participants of the project. The rating team will attend a distanced training course taught by WHO experts on how to adopt the JERI on assessing caregiver-child gameplay videos. There will be 5 sessions (about 2 hours) in the whole training course. The trainers from WHO will ensure that the raters will be qualified to use JERI in rating the videos.

To ensure the reliability of the coding scheme, one-third of the videos will be blindly sent to two raters for rating, and inter-rater reliability of each item in the JERI will be calculated.

## Date Analysis

Two sources of data will be collected: (1) data from the online survey platform, (2) data collected from the JERI ratings video recording caregiver-child interactions. For the variables which presented as continuous or count data, the between group mean differences, the standard deviation, the range and the possible range (as provided in the instrument), at different time points of assessment will be reported. Given the intent-to-treat design, all caregiver-children dyads’ data will be contained in the original assignment group, including those who dropped out before the end of the study, and all will take the assessment at any of the three timepoints as possible. All the outcome measurements with continuous variables (i.e., JERI, GHQ12, ECBI, etc.) will be analyzed using linear mixed models (LMM) with random intercept and slope parameters, where appropriate, examining the effect of treatment assignment (treatment group versus wait-list-control group), time points of assessment (T1, T2 and T3), and their interaction on outcome. The reason of using the LMM is because of its full-information loaded characteristic which can involve information from all randomized (intent-to-treat) participants, including those with only partial data owing to drop-out or other reasons (Unnebrink & Windeler, 2001). LMM are advantageous compared to repeated measures ANCOVA in that they accommodate data of missing time points, hence utilize all available data, and therefore can be considered in our intent-to-treat models. One’s latest assessments before dropout or completion of the programme will be used in the model. The assessment at T2 and its interaction with group and time are the primary interest to test our hypotheses. The LMM analysis will be performed by R package lme4 and glmmTMB.

## Ethical Considerations

Before the commencement of the study, ethical approval has already been obtained from the Human Research Ethics Committee, the University of Hong Kong. There is no foreseeable significant psychological distress and any other hazards entailed by the study procedures.

### Plan for Obtaining Informed Consent

Written consent will be obtained from all the participants before completing the baseline assessment. The participation is entirely voluntary; every participant maintains every right to withdraw from the study. For the caregivers, they will be further reassured that their participation will not affect the health care and/or social services that the families and their children are currently receiving. The text in the Informed Consent Form will be read to all potential participants (i.e., caregivers) and they will have an opportunity to ask questions and express concerns (e.g., the purpose of video-recording). The minimal time given to the caregivers for consideration after explanation will be 15 to 20 minutes. However, the caregivers can take as much time as needed to consider taking part of the study. In addition, the caregivers will be informed verbally about future publications of the study in the scientific literature. All the information obtained will be as confidential.

### Anonymousness and Confidentiality

All data collected will be kept confidential and be used for research purposes only. Each participant will be assigned a unique identification code (ID code). ID codes instead of names of participants will be used and thus, identity of participants would not be disclosed to unauthorized persons. Moreover, the ID codes with names will be stored separately in documents from the collected data.

Data with identifiable information will be kept in a locked cabinet, which can only be accessed by the research team. All personal data and video-recorded files will be stored in password-protected files and encrypted. No data will be stored on personally-owned personal computers or portable storage devices. No personal identifiable information will be reported in any of the reports or publications.

Concerning the video-records made during the home-visit, all the video-records will be uploaded by facilitators to an online platform developed by IT staffs of Faculty of Social Sciences, the University of Hong Kong. Only researchers have the rights to assess the online platform which is consistently monitored by IT staffs of Faculty of Social Sciences. Therefore, the privacy of participants can be protected.

## Roles and Responsibilities

### Role of Coordinating Team

The Family Support Team of the JC A-Connect project is subordinated the Faculty of Social Science, The University of Hong Kong. The team is responsible for delivering the clinical services filling the syllabus and standard of WHO-CST, training trainers to deliver the WHO-CST programme at community level, collecting and analyzing data for the evaluation of WHO-CST programme in Hong Kong.

## Limitation

The aim of this study is to evaluate the implementation of the WHO-CST programme in Hong Kong by measurable and scientific method. This research will involve human participants who are supposed to be caregivers of ASD children (or suspected ASD children). As mentioned at the introduction, parents of ASD children are so stressful in Hong Kong, also it is a common practice in Hong Kong for both parents of a children having their own jobs. There will be possibility that caregivers may drop out from the RCT due to emotional or personal (e.g. time management) reasons. Although the Team expected 10% dropout rate in the proposal, the actual rate is not so anticipatable and may be higher than expected. The Team will try to recruit participants more than the suggested number to prevent the failure of research study causing by the dropout problem.

Another limitation is about the changed mode of the WHO-CST delivered. The original WHO-CST should be delivered by face-to-face lectures and tutorial, and there should be three session of actual home-visit for trainers to provide guidance on parent-child interaction. Under the pandemic of COVID-19, face-to-face lessons are sometimes prohibited by the government or the university. Therefore, our WHO-CST lessons will be probably delivered in the form of distanced courses (e.g. by ZOOM), and the home-visits will become sessions situated in the University of Hong Kong. Our evaluation results and the effect of training can only represent an “on-line delivered version” of WHO-CST, but the traditional face-to-face basis WHO-CST.

## Data Availability

The data will be available at July 2020. The use of data will be subject to the agreement with the WHO side since a team of WHO staff has been involved in the development of this study.

## Footnotes

3 The nine countries were China, Korea, India, Lebanon, Bangladesh, Iran, Israeli, Nepal and Sri Lanka.

